# The potential causal association of systemic lupus erythematosus with congestive heart failure in the East Asian population: A two-sample Mendelian randomization study

**DOI:** 10.1101/2022.11.17.22282308

**Authors:** Euijun Song

## Abstract

**Objective:** Systemic lupus erythematosus (SLE) is a systemic autoimmune disease that increases the risk of cardiovascular disease, but the causal relationship has remained unknown in the East Asian population. We aim to determine the causal relationship between SLE and congestive heart failure (CHF) in the East Asian population.

**Methods:** We conducted a two-sample Mendelian randomization (MR) study to examine the potential causal association of SLE with CHF, using the East Asian genome-wide association study datasets for SLE (4,222 cases, 8,431 controls) and CHF (9,413 cases, 203,040 controls).

**Results:** The MR analysis showed that genetic susceptibility to SLE was associated with the increased risk of CHF (odds ratio [OR] 1.032, 95% confidence interval [CI] 1.004–1.061, P=0.023). After excluding the human leukocyte antigen (HLA) regions, SLE was also associated with a higher risk of CHF (OR 1.034, 95% CI 1.007–1.062, P=0.015). The multiple MR sensitivity analyses confirmed that this potential causal association was unlikely to be biased by horizontal pleiotropy.

**Conclusions:** The findings of this MR study suggest that SLE potentially increases the risk of CHF in the East Asian population. Genetic predisposition to SLE may play a significant role in developing CHF in the East Asian population.

**What is already known on this topic**

- Patients with systemic lupus erythematosus (SLE) have a higher risk of congestive heart failure (CHF) in the European and East Asian populations.

**What this study adds**

- The Mendelian randomization analysis suggests that genetic susceptibility to SLE was associated with a higher risk of CHF in the East Asian population.
- Non-human leukocyte antigen (HLA) variants associated with SLE were also associated with the increased risk of CHF.

**How this study might affect research, practice, or policy**

- Genetic predisposition to SLE may be useful to understand CHF risk stratification and management in the East Asian population.

## 1. Introduction

Systemic lupus erythematosus (SLE) is a systemic autoimmune disease that can affect a variety of organs and systems in the human body, leading to rash, arthritis, lupus nephritis, anemia, and serositis. Patients with SLE have diverse autoantibodies against double-stranded DNA, cardiolipin, and ribonucleoproteins. Cardiovascular disease and infections are the leading causes of death in patients with SLE (1). Cardiovascular manifestation and complications of SLE include pericarditis, coronary artery disease, arrhythmias, valvular heart disease, and heart failure (2). Several retrospective cohort analyses and Mendelian randomization (MR) studies demonstrated that SLE increases the risk of coronary artery disease and heart failure, and worsens cardiovascular outcomes (3-6). However, most studies have been conducted in the European-dominant population, and the underlying mechanisms remain unclear.

SLE and cardiovascular disease are complex diseases, and genetic predisposition plays an important role in both diseases (5). Here, we conduct a two-sample MR study to examine the potential causal association of SLE with congestive heart failure (CHF) in the East Asian population. We use large-scale summary-level genome-wide association study (GWAS) datasets for SLE and CHF in the East Asian population, and perform multiple MR methods to determine the potential causal effects of SLE on CHF. Since genetic variants within the human leukocyte antigen (HLA) region are not well understood and highly pleiotropic, we also performed our MR analysis after excluding the extended HLA region.

## 2. Methods

### 2.1. GWAS datasets for SLE and CHF

To perform the two-sample MR, we first recruited publicly available GWAS summary-level datasets for SLE and CHF in the East Asian population. For SLE, we used a Chinese GWAS summary-level dataset of 4,222 SLE cases and 8,431 controls from Wang et al (7). For CHF, we used the Biobank Japan GWAS summary statistics of 9,413 CHF cases and 203,040 controls (available from https://gwas.mrcieu.ac.uk/, dataset ID: bbj-a-109, accessed on Nov 6, 2022).

### 2.2. MR and statistical analysis

We constructed genetic instruments for SLE by extracting genome-wide significant variants (P<5×10^−8^) that were in low linkage disequilibrium (r^2^<0.001). The clumping was performed using the *clump_data* function of the TwoSampleMR package based on the East Asian 1000 Genomes Project reference panel. We only considered non-palindromic variants in both SLE and CHF GWAS datasets. The instrument strength was evaluated using the F-statistic as 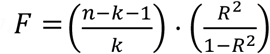, where *n* is the sample size, *k* is the number of the genetic variants, and *R*^2^ is the proportion of variance explained by the genetic variants. The F-statistic greater than 10 indicates valid instruments. To evaluate the putative causal effect, we performed the inverse- variance weighted (IVW), weighted median, maximum likelihood, MR-RAPS (8), MR-PRESSO (9), and radial MR (10) analyses. The heterogeneity between SNPs was evaluated by Cochran’s Q test. The MR Steiger directionality test was also performed to determine the directionality of causality (11). To examine potential horizontal pleiotropy, we used the MR-Egger intercept test (12) and MR-PRESSO method. We further performed the leave-one-out sensitivity analysis and radial MR to identify potential outlying variants. The P-value<0.05 was considered statistically significant. All MR analyses were performed with the TwoSampleMR, RadialMR, MR.RAPS, and MRPRESSO packages of R (version 4.1.2).

## 3. Results

### 3.1 The causal association of SLE with CHF

We used 30 genome-wide significant variants (P<5×10^−8^) as SLE-associated instrumental variables (Table S1). The genetic instruments for SLE have an F-statistic of 81.8, indicating no significant weak instrument bias in our MR analysis. We performed a two-sample MR analysis to examine the putative causal effects of SLE on CHF in the East Asian population. As highlighted in Table 1 (left column), the IVW method showed that SLE potentially increased the risk of CHF (odds ratio [OR] 1.032, 95% confidence interval [CI] 1.004–1.061, P=0.023). There was no evidence of heterogeneity (Cochran’s Q-statistic=33.122, P=0.273). The weighted median, maximum likelihood, radial IVW, and MR-RAPS methods also showed directionally similar results (Table 1). Moreover, the MR Steiger directionality test confirmed the direction of the causality (P<0.001).

**Table 1.** Mendelian randomization (MR) analyses between systemic lupus erythematosus (SLE) and congestive heart failure (CHF).

|  | Whole SLE-associated SNP on CHF |  |  | Non-HLA SLE-associated SNP on CHF |  |  |
| --- | --- | --- | --- | --- | --- | --- |
|  | OR (95% CI) | P-value | NSNP | OR (95% CI) | P-value | NSNP |
| IVW | 1.032 (1.004–1.061) | 0.023 | 30 | 1.034 (1.007–1.062) | 0.015 | 27 |
| Weighted median | 1.024 (0.985–1.064) | 0.233 | 30 | 1.024 (0.986–1.063) | 0.223 | 27 |
| Maximum likelihood | 1.033 (1.006–1.060) | 0.015 | 30 | 1.034 (1.007–1.063) | 0.013 | 27 |
| IVW (Radial) | 1.032 (1.004–1.061)* | 0.023 | 30 | 1.034 (1.007–1.062)** | 0.015 | 27 |
| MR-RAPS | 1.032 (1.003–1.061) | 0.029 | 30 | 1.032 (1.004–1.062) | 0.024 | 27 |
| MR Steiger directionality | - | <0.001 | 30 | - | <0.001 | 27 |
| MR-PRESSO<sup>†</sup> | - | 0.451 | 30 | - | 0.689 | 27 |
| MR-Egger intercept<sup>‡</sup> | 0.01067 (0.01072) | 0.328 | 30 | 0.00930 (0.01051) | 0.385 | 27 |

OR, odds ratio; CI, confidence interval; SNP, single nucleotide polymorphism; NSNP, the number of SNPs;
IVW, inverse-variance weighted method; HLA, human leukocyte antigen.
\*The outlying variants (rs143821594 and rs4930642) were detected.
\*\*The outlying variant (rs4930642) was detected.
<sup>†</sup>No outlying pleiotropic variants were detected.
<sup>‡</sup>The Egger intercept value and SE are presented. There was no evidence of significant horizontal pleiotropy.

To examine potential horizontal pleiotropy, which could bias the MR analysis, we performed the MR-Egger intercept test and MR-PRESSO method. The MR-Egger intercept test showed no evidence of significant horizontal pleiotropy (intercept=0.01067, P=0.328). The MR-PRESSO global test further indicated no significant pleiotropy with no outlying genetic variants (P=0.451). The removing individual variants did not substantially alter the estimated causal effect (Figure S1A). The radial MR method identified the outlying variants (rs143821594 and rs4930642); however, the excluding that outlier also led to the concordant MR result (OR 1.032, 95% CI 1.004–1.061, P=0.023; Figure S2A). Additionally, the MR-RAPS was used to test whether SLE affects CHF through weak instruments (OR 1.032, 95% CI 1.003–1.061, P=0.029). Multiple sensitivity analyses indicate no significant evidence for pleiotropic effects in our MR analysis.

### 3.2. Effects of non-HLA SLE-associated variants on CHF

Since GWAS loci within the HLA region are difficult to finely map and are highly pleiotropic to complex diseases, we repeated our MR analysis after excluding the extended HLA region (chromosome 6: 27–34 Mb). Three genetic variants (rs2524058, rs6941112, and rs143821594) were excluded. Using IVW, we found that SLE was causally associated with the risk of CHF (OR 1.034, 95% CI 1.007–1.062, P=0.015). The weighted median, maximum likelihood, radial IVW, and MR-RAPS methods showed directionally similar estimates (Table 1, right column). The MR-Egger intercept test (intercept=0.00930, P=0.385) and MR-PRESSO global test indicated no significant evidence of horizontal pleiotropy (P=0.689). The leave-one-out sensitivity analysis and the radial plot are presented in Figure S1B and Figure S2B, respectively. Furthermore, we also found that SLE was causally associated with CHF after excluding the entire chromosome 6 (OR 1.037, 95% CI 1.004–1.072, P=0.030; Table S2). The overall results are concordant with those acquired from the whole SLE-associated variants.

## 4. Discussion

The causal relationship between SLE and CHF in the East Asian population has been largely unknown. In the present MR study, we discovered that SLE potentially increased the risk of CHF in the East Asian population. The non-HLA SLE-associated variants were also causally associated with CHF. The multiple MR sensitivity analyses confirmed that this potential causal association was unlikely to be biased by horizontal pleiotropy, indicating the robustness of effect estimates.

In the US retrospective studies, SLE was associated with a 2.7–3.17-fold increase in the risk of CHF (3, 4). Recently, European-dominant MR studies also revealed that SLE potentially increases the risk of CHF, coronary artery disease, ischemic stroke, and venous thromboembolism (5, 6). A Korean nationwide study found that patients with SLE had a 4.6-fold higher risk of CHF (13). In our MR study, the effect size of SLE on CHF (OR 1.032, 95% CI 1.004– 1.061) was relatively smaller compared to previous retrospective studies. Therefore, developing CHF in patients with SLE may be attributed to other environmental factors and SLE medications, such as hydroxychloroquine and steroid, as well as the genetic predisposition for SLE *per se*.

Indeed, hydroxychloroquine and steroid, commonly used medications in SLE, increases the risk of diabetes mellitus, dyslipidemia, and cardiomyopathy (14, 15). These drug-induced risk factors for cardiovascular diseases may lead to bias in retrospective cohort studies. In contrast, we demonstrated that genetically predisposed SLE was associated with CHF in the East Asian population using the MR analysis. Thus, MR approaches could identify potential causal associations without conducting prospective clinical trials.

There are several limitations of this study. There is limited availability of large-scale SLE GWAS datasets, especially for the East Asian population, restricting statistical power underlying the polygenic pathobiology of SLE. In our analysis, the weighted median method did not show significant results; however, IVW is more statistically powerful than other MR methods. We also did not perform replication analysis in other East Asian datasets, though large-sale SLE GWAS datasets are not currently available. The linkage disequilibrium score regression and pathway/network analyses would uncover potential pathobiological connections between SLE and CHF.

## Data Availability

All data produced in the present study are available upon reasonable request to the authors.

## Acknowledgements

This research received no external funding. In the present work, we only used publicly available GWAS summary statistics datasets. The author would like to thank Dr. Cheng for his kind support.

---

**Figure S1.**
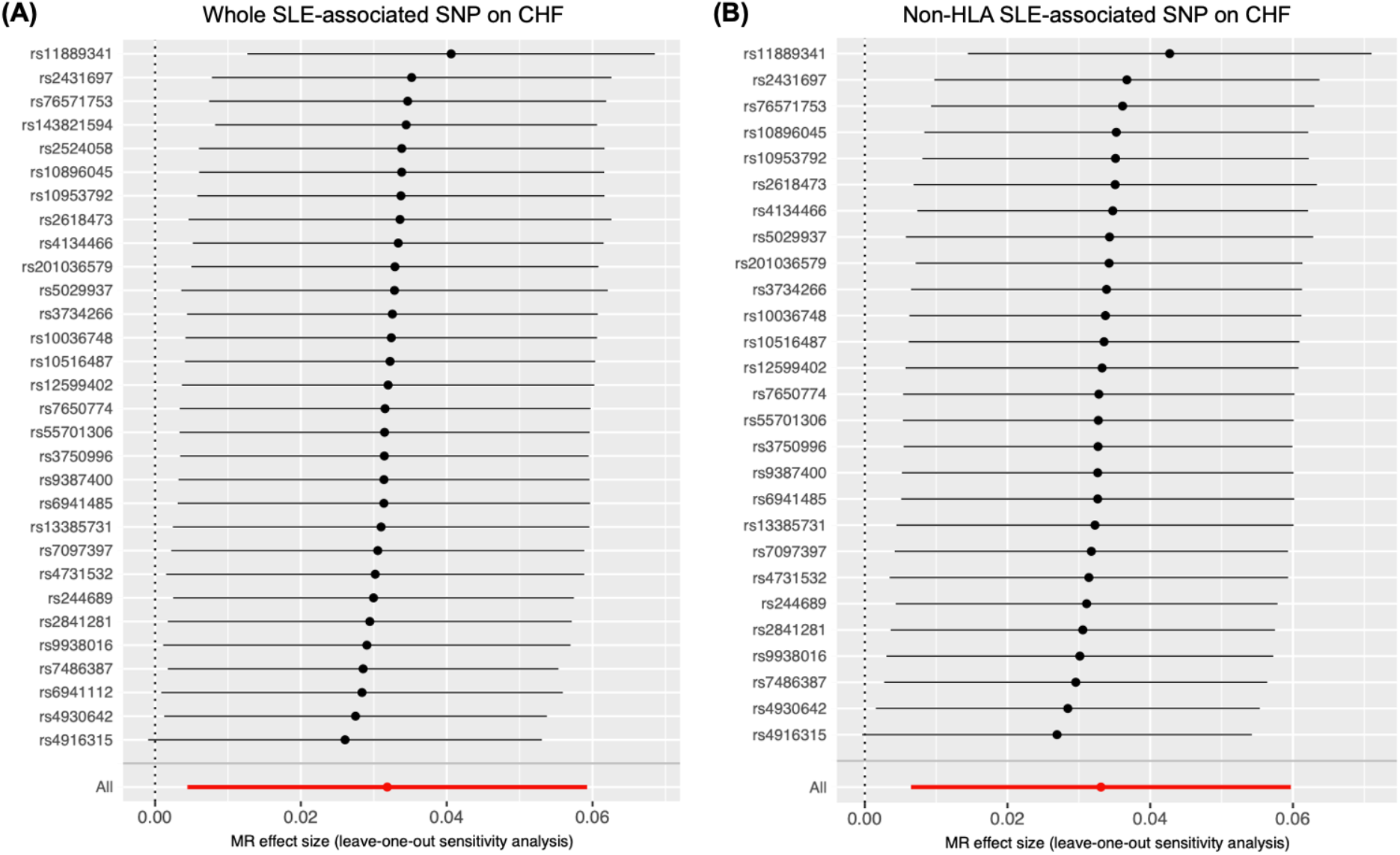
The leave-one-out sensitivity analysis for the causal association of systemic lupus erythematosus with congestive heart failure using the inverse-variance weighted method. (A) The whole genetic variants associated with SLE were used. (B) The only non-HLA genetic variants were used. SLE, systemic lupus erythematosus; CHF, congestive heart failure; HLA, human leukocyte antigen.

**Figure S2.**
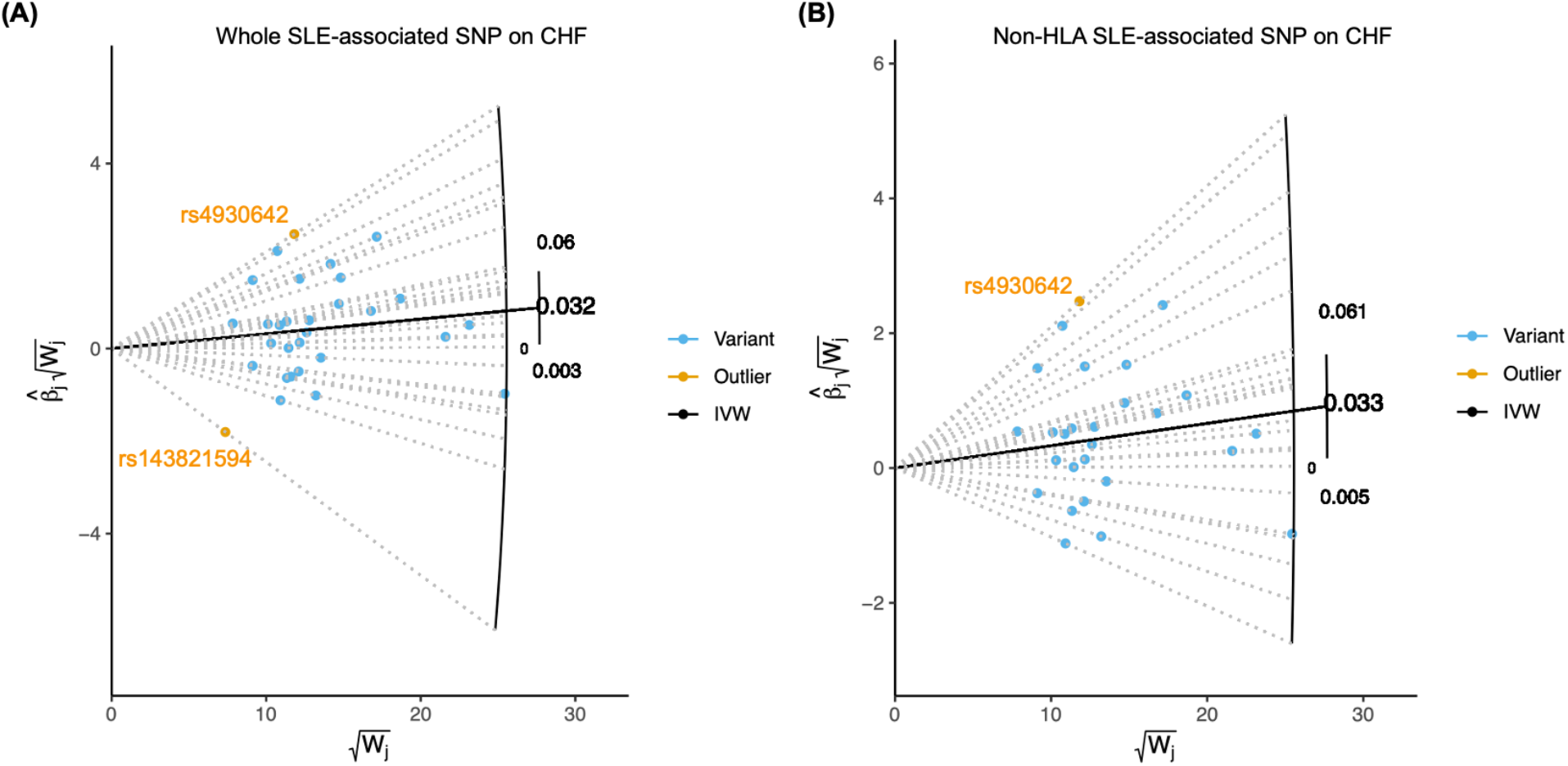
The radial Mendelian randomization plot. Blue and orange dots indicate valid genetic variants and the outliers (rs143821594 and rs4930642), respectively. The estimated effect sizes are presented. (A) The whole genetic variants associated with SLE were used. (B) The only non-HLA genetic variants were used. IVW, inverse-variance weighted method; SLE, systemic lupus erythematosus; CHF, congestive heart failure; HLA, human leukocyte antigen.

**Table S1.** Genetic instruments for systemic lupus erythematosus in the East Asian population (P<5×10^−8^).

| SNP | Position (GRCh37) | EA/OA | Beta (SE) | P-value |
| --- | --- | --- | --- | --- |
| rs201036579 | chr1:157499390 | C/T | -0.288 (0.053) | 4.85E-08 |
| rs4916315 | chr1:173215372 | T/C | 0.336 (0.031) | 4.12E-27 |
| rs13385731 | chr2:33701890 | C/T | -0.365 (0.042) | 4.19E-18 |
| rs11889341 | chr2:191943742 | T/C | 0.416 (0.03) | 3.33E-45 |
| rs7650774 | chr3:119205050 | C/T | -0.18 (0.031) | 4.07E-09 |
| rs10516487 | chr4:102751076 | A/G | -0.255 (0.04) | 1.64E-10 |
| rs244689 | chr5:133422816 | G/A | -0.16 (0.029) | 4.29E-08 |
| rs10036748 | chr5:150458146 | T/C | 0.202 (0.034) | 2.29E-09 |
| rs2431697 | chr5:159879978 | C/T | -0.254 (0.045) | 1.84E-08 |
| rs6941485 | chr6:250619 | G/A | 0.203 (0.031) | 9.53E-11 |
| rs2524058* | chr6:31251569 | G/A | -0.186 (0.03) | 5.31E-10 |
| rs6941112* | chr6:31946614 | A/G | 0.222 (0.033) | 1.01E-11 |
| rs143821594* | chr6:32204017 | C/T | -0.495 (0.055) | 3.08E-19 |
| rs3734266 | chr6:34823187 | C/T | 0.334 (0.04) | 6.09E-17 |
| rs4134466 | chr6:106577368 | G/A | -0.214 (0.031) | 3.09E-12 |
| rs9387400 | chr6:116694120 | A/C | -0.297 (0.054) | 3.14E-08 |
| rs5029937 | chr6:138195151 | T/G | 0.684 (0.068) | 4.32E-24 |
| rs76571753 | chr7:73974915 | T/G | 0.279 (0.041) | 1.46E-11 |
| rs10953792 | chr7:75172937 | C/T | 0.199 (0.032) | 4.69E-10 |
| rs4731532 | chr7:128572766 | A/G | 0.372 (0.033) | 8.15E-29 |
| rs2618473 | chr8:11344127 | T/C | 0.359 (0.034) | 2.89E-26 |
| rs7097397 | chr10:50025396 | A/G | -0.239 (0.031) | 1.32E-14 |
| rs3750996 | chr11:4113200 | G/A | -0.204 (0.037) | 4.91E-08 |
| rs10896045 | chr11:65555524 | G/A | -0.173 (0.029) | 1.78E-09 |
| rs4930642 | chr11:68816370 | G/A | -0.234 (0.034) | 1.10E-11 |
| rs7486387 | chr12:12866911 | A/G | -0.189 (0.031) | 1.98E-09 |
| rs2841281 | chr14:105394669 | T/C | 0.193 (0.029) | 3.56E-11 |
| rs12599402 | chr16:11189888 | C/T | -0.198 (0.029) | 1.02E-11 |
| rs9938016 | chr16:86002524 | T/C | -0.397 (0.059) | 1.78E-11 |
| rs55701306 | chr17:16842447 | T/C | 0.166 (0.029) | 1.38E-08 |

SNP, single nucleotide polymorphism; EA, effect allele; OA, other allele; SE, standard error.
\*SNPs located in the extended HLA region (chromosome 6: 27–34 Mb).

**Table S2.**
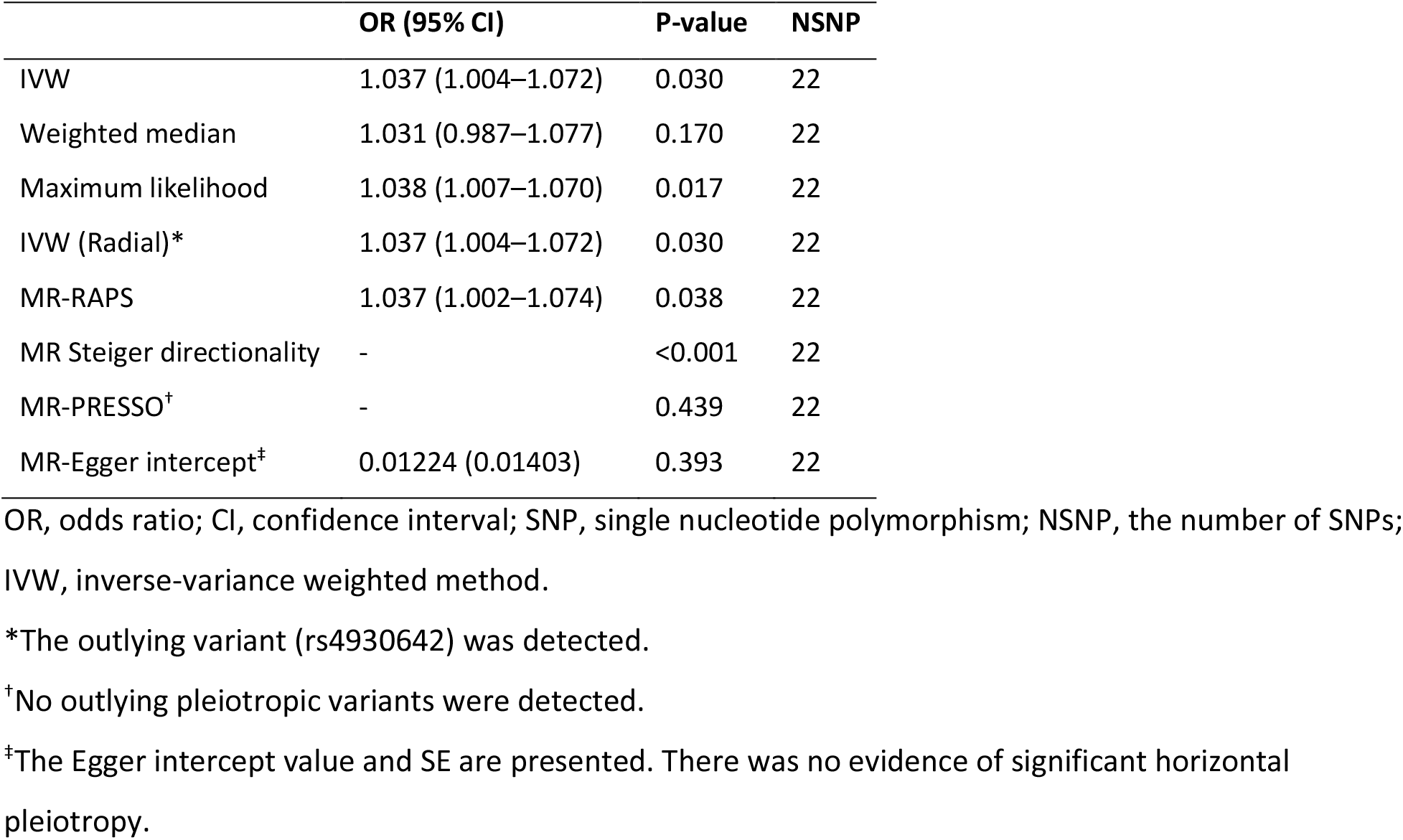
Mendelian randomization (MR) analyses between systemic lupus erythematosus (SLE) and congestive heart failure (CHF) after excluding the entire chromosome 6.

|  | OR (95% CI) | P-value | NSNP |
| --- | --- | --- | --- |
| IVW | 1.037 (1.004–1.072) | 0.030 | 22 |
| Weighted median | 1.031 (0.987–1.077) | 0.170 | 22 |
| Maximum likelihood | 1.038 (1.007–1.070) | 0.017 | 22 |
| IVW (Radial)* | 1.037 (1.004–1.072) | 0.030 | 22 |
| MR-RAPS | 1.037 (1.002–1.074) | 0.038 | 22 |
| MR Steiger directionality | - | <0.001 | 22 |
| MR-PRESSO<sup>†</sup> | - | 0.439 | 22 |
| MR-Egger intercept<sup>‡</sup> | 0.01224 (0.01403) | 0.393 | 22 |

OR, odds ratio; CI, confidence interval; SNP, single nucleotide polymorphism; NSNP, the number of SNPs; IVW, inverse-variance weighted method.
\*The outlying variant (rs4930642) was detected.
<sup>†</sup>No outlying pleiotropic variants were detected.
<sup>‡</sup>The Egger intercept value and SE are presented. There was no evidence of significant horizontal pleiotropy.

